# Comparisons Between GPS-based and Self-reported Life-space Mobility in Older Adults

**DOI:** 10.1101/2022.09.19.22280116

**Authors:** Chen Bai, Ruben Zapata, Yashaswi Karnati, Emily Smail, Alexandra M. Hajduk, Thomas M. Gill, Sanjay Ranka, Todd M. Manini, Mamoun T. Mardini

## Abstract

Assessments of Life-space Mobility (LSM) evaluate the locations of movement and their frequency over a period of time to understand mobility patterns. Advancements in and miniaturization of GPS sensors in mobile devices like smartwatches could facilitate objective and high-resolution assessment of life-space mobility. The purpose of this study was to compare self-reported measures to GPS-based LSM extracted from 27 participants (44.4% female, aged 65+ years) who wore a smartwatch for 1-2 weeks at two different site locations (Connecticut and Florida). GPS features (e.g., excursion size/span) were compared to self-reported LSM with and without an indicator for needing assistance. Although correlations between self-reported measures and GPS-based LSM were positive, none were statistically significant. The correlations improved slightly when needing assistance was included, but statistical significance was achieved only for excursion size (r=0.40, P=0.04). The poor correlations between GPS-based and self-reported indicators suggest that they capture different dimensions of life-space mobility.

## Introduction

The maintenance of mobility is fundamental for older adults to stay independent and autonomous^1^. Broadly defined, mobility is the ability to move independently around the environment^1,2^. Previous studies have shown that mobility performance is associated with frailty, cognitive decline, and all-cause mortality^2–6^ and is predictive for the onset of functional dependence^7^. Mobility function can be assessed through self-reports of ability to perform specific activities (e.g. walking a block) and the location of travel inside and outside the community with and without transportation. Objective indicators include accelerometers that measure steps or global positioning system (GPS) that assess travel between distant locations^1,8^.

Life-space mobility (LSM) integrates the measurement of area, frequency, and independence of individuals’ movement^9,10^. LSM is most commonly measured using self-reported questionnaires^9,11–14^ and is associated with health outcomes such as quality of life, falls, hospital readmission, and mortality^15–21^. May et al.^12^ introduced the first assessment of LSM in 1985 by asking the participants to keep a diary of the zones (5 pre-defined zones in total) in which they moved during the day. The Life-space Questionnaire, introduced by Stalvey et al.^13^, includes 9 (Yes/No) questions that cover several levels of movement between rooms in the house up to traveling outside of the United States over a period of time. The most widely used questionnaire to assess LSM in community-dwelling adults is the University of Alabama at Birmingham Life-Space Assessment (UAB-LSA)^11^. The UAB-LSA measures not only the life-space of the participants, but also the frequency and need for assistance. However, self-reported assessment of LSM adds additional burden to participants and leads to recall and social desirability bias^9,22^.

GPS devices offer researchers a more convenient and flexible approach to collect high resolution geographical data for extended periods of time. GPS devices have been used to measure the activity space and life-space mobility in older adults^23–27^. Wearable devices, such as smartwatches, that are coupled with a GPS receiver provide an unobtrusive approach to continuously collect GPS data. According to the International Data Corporation, the shipment of wearable devices will reach 637.1 million units in 2024^28^. Smartwatches have been used in many studies to measure mobility and record travel trajectories in older adults^27,29–32^. However, the convergent validity between the GPS features collected through smartwatches and self-reported responses obtained from an LSM questionnaire has been rarely examined^26,33^. Elucidating the relationship between the GPS features and self-reported responses provides a basis for better understanding the dimensions of LSM.

In this study, we compared the GPS features extracted from the raw GPS coordinates collected from a custom-designed smartwatch platform called Real-time Online Assessment and Mobility Monitor (ROAMM)^34^ to self-reported LSM using the UAB-LSA questionnaire. (Figure 1) provides an overview of the design of the study.

**Figure 1.**
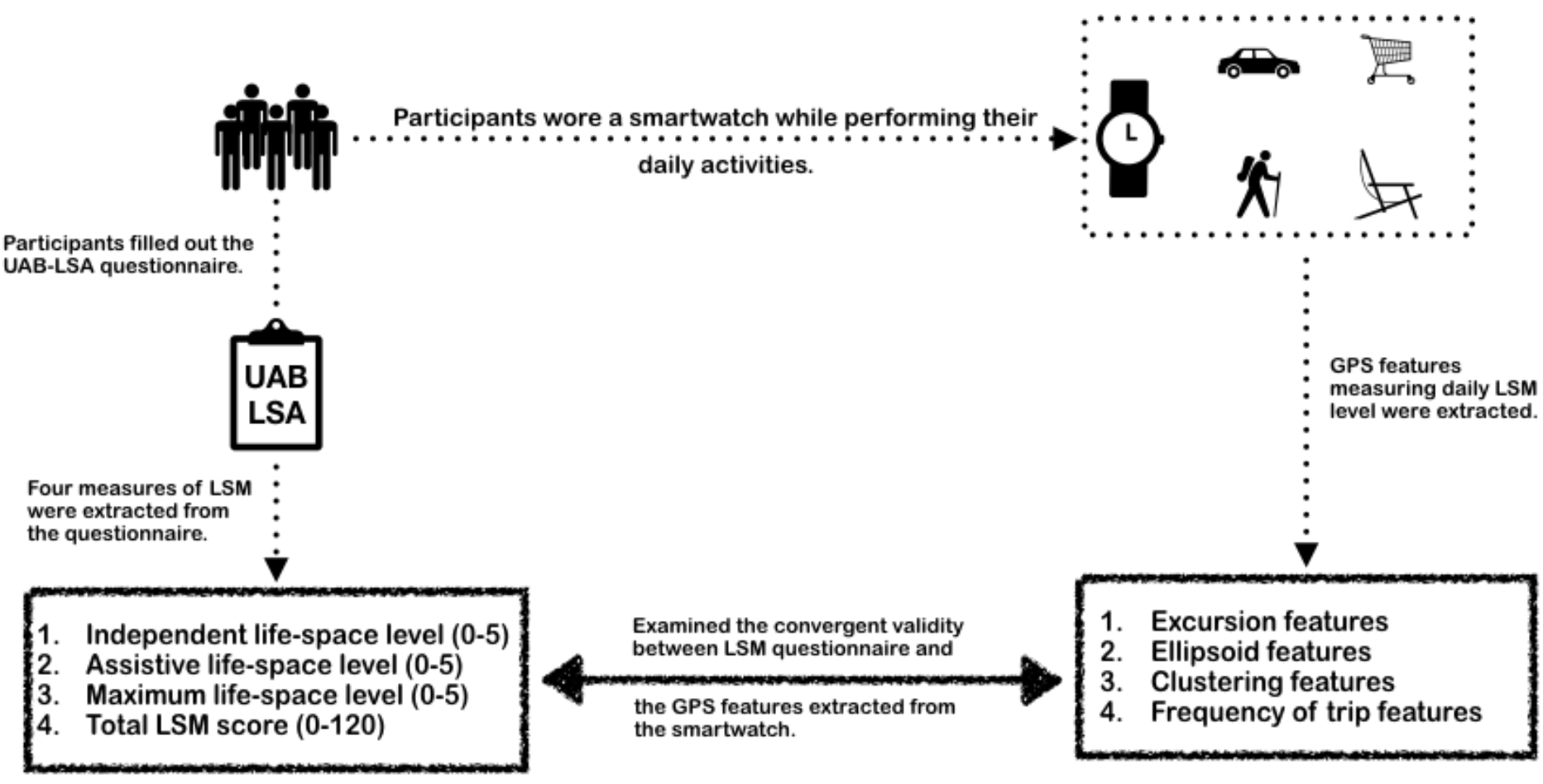
An overview of the design of the study.

## Methods

### Participants

This study was approved by the University of Florida (UF) institutional review board and Yale University (Yale) institutional review board. Written informed consent was obtained from all participants. Participants were community dwelling adults age 65+ years who could read and speak English and were willing to undergo all testing procedures. Participants were excluded from the study if they were unable to provide informed consent, lived in a nursing home, used a wheelchair, had major ADL disability (unable to feed, dress, bath, use the toilet, or transfer), had severe sensory deficit (severe hearing loss, speech disorder or blindness), had a planned surgical procedure or hospitalization in the next 12 month, or had any other significant conditions or health behaviors discovered during medical screening that would impact safety and/or compliance to the protocol (e.g., psychiatric disorder – bipolar, schizophrenia, and dementia). Participants’ demographics were recorded and their lower extremity function was assessed using Short Physical Performance Battery (SPPB)^35^ during their in-person visits. GPS data were collected at two sites — 21 participants from Gainesville, Florida and 13 participants from New Haven, Connecticut. We excluded participants (n=7) who did not have complete data, leaving a total of 27 participants for this analysis.

### GPS Data Collection, Processing, and Feature Extraction

Participants were asked to wear an Apple Watch SE or a Samsung Galaxy 3 during the waking day for approximately 1-2 weeks. GPS coordinates were sampled every fifteen minutes throughout the day. Days that had less than10 hours of data collection were excluded. Ten GPS features were extracted based on our previous work^27^ and were summarized into four categories: excursion, ellipsoid, clustering, and trip frequency. (Table 1) provides definitions of the extracted features, and (Figure 2) provides a graphical illustration of the ellipse encompassing all the GPS coordinates of a participant during a 1-day time frame. The extraction of the GPS features depends on the participant’s home address. Unfortunately, not all home addresses could be shared because of privacy laws. For those without a home address (n = 9), the time derivative of each GPS coordinate was calculated and GPS coordinates were labelled as either stationary (time derivative < 1 km/h) or moving. An adaptive *k* -means algorithm was used to cluster the stationary GPS coordinates. The centroid of the cluster containing the highest number of GPS coordinates was selected as the estimated home address for the participant for that day. In this way, each participant had one estimated home address for each day. The final home address was subsequently determined to be the location with the largest number of identified clusters. For home address estimation, we only considered GPS coordinates at night-time to remove the influence of activities during the daytime. Using data collected from UF, we validated and tuned our estimation algorithm by evaluating centroids variation across days for each participant. 75% of the home addresses were accurately estimated.

**Table 1.**
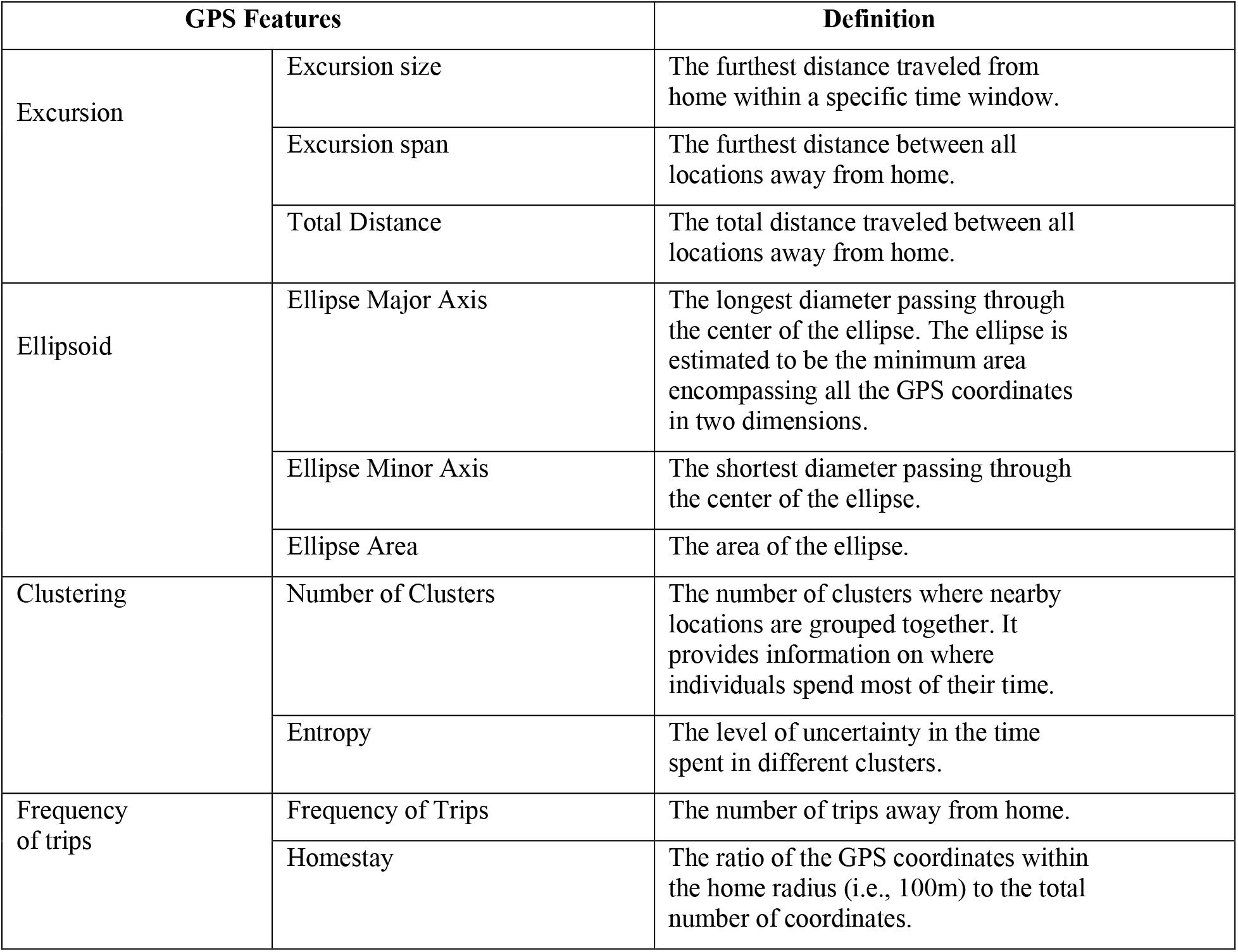
Summary of definitions of the extracted GPS features.

**Figure 2.**
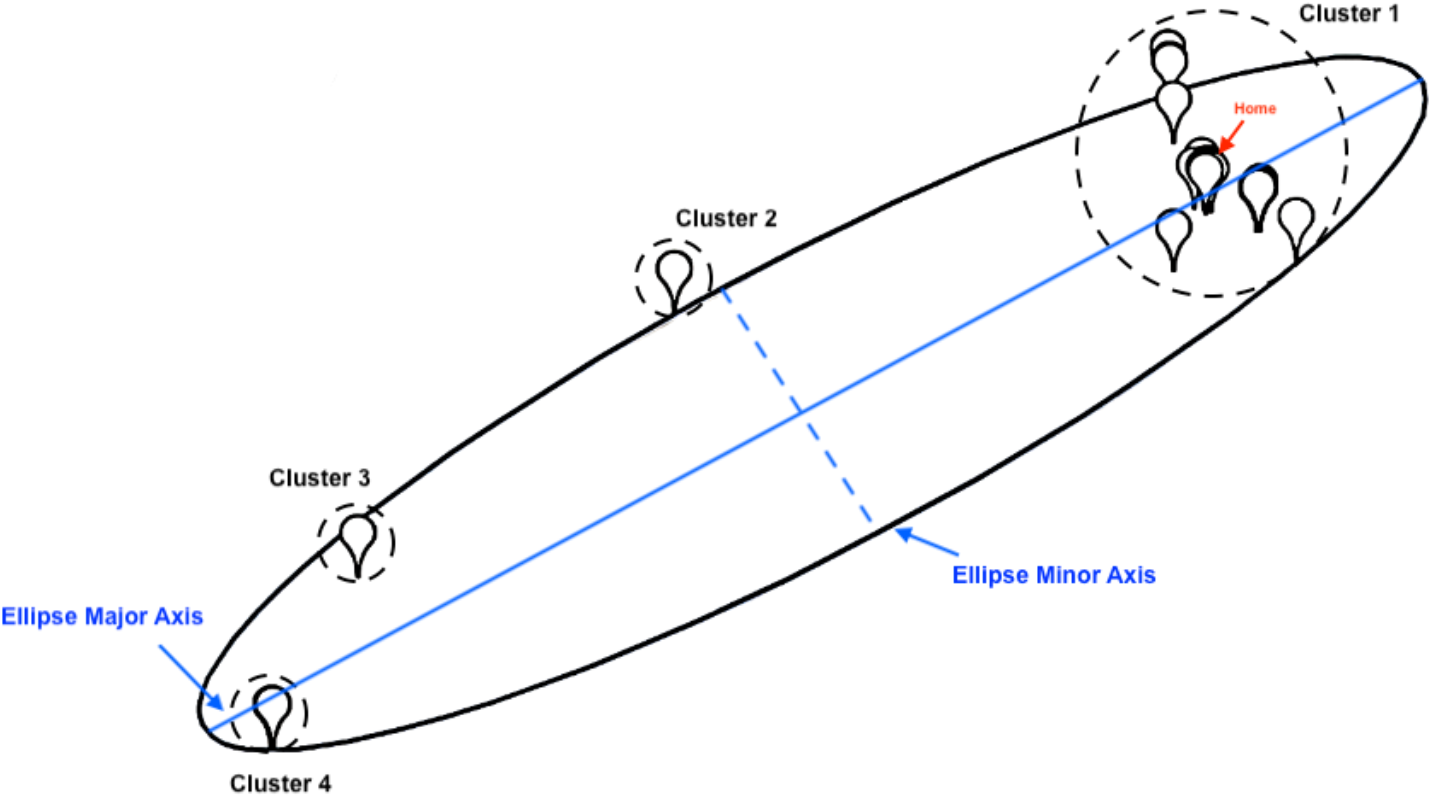
An illustration of the ellipse encompassing all the GPS coordinates for a participant during a 1-day time frame. The ellipse in black is estimated based on all the GPS coordinates. The solid blue line passing through the center of the ellipse is the major axis. The blue dash line which is orthogonal to the major axis is the minor axis. The four clusters identified are shown on the map, each containing a set of GPS coordinates. The red arrow points to the home of the participant.

### UAB Life-Space Assessment

Participants were asked to fill out the University of Alabama at Birmingham Life-Space Assessment^9^ (UAB-LSA) during their in-person visit. The UAB-LSA is the most widely used instrument to measure life-space among community-dwelling adults. The UAB-LSA measures the frequency of five life-space levels (1: other rooms at home besides the room where individuals sleep, 2: an area outside individuals’ home, 3: places within individuals’ neighborhood, 4: places outside individuals’ neighborhood, and 5: places outside of individuals’ town) that a person has mobilized during the prior 4-week period, while considering the use of assistance. Each life-space level is assigned an index from 1 to 5 depending on the distance from the person’s bedroom. A sub-score for each life-space level is calculated by multiplying the index, the frequency (1 to 4, from less than once per week to daily), and the use of assistance (1: personal assistance, 1.5: equipment only, and 2: no equipment or personal assistance). The composite LSA score, ranging from 0 to 120, is calculated as the sum of sub-scores from five life-space levels.

Additional measures of LSM can be derived based on the highest level of life-space without considering the frequency of actions^11^. These measures include 1) independent life-space level (1-5), the highest level of life-space achieved without any assistance; 2) assistive life-space level (1-5), the highest level of life-space achieved without help from another person, even if the equipment was used; and 3) maximal life-space (1-5), the highest level of life-space achieved regardless of the use of personal or equipment assistance. The composite LSA score, independent, assistive, and maximal life-space for each participant were calculated and compared with GPS features.

### Statistical Analysis

The excursion size, excursion span, total distance, ellipse major axis, ellipse minor axis, and the ellipse area were transformed into log values to avoid skewness. GPS features were aggregated for each participant and the average value of each feature for each day was taken to represent the daily LSM. Spearman’s correlation coefficients between the average value of each GPS feature and each measure extracted from the UAB-LSA were calculated. The significance level was set at P < 0.05.

## Results

(Table 2) shows a summary of measures of LSM derived from the UAB-LSA in our participants. The mean LSA score was 79. More than half of the participants had an independent life-space level of 5 (were able to go out of town without assistance). Two participants had an independent life-space level of 1 (were not able to go outside of home without any assistance). All participants had an assistive life-space level greater than or equal to 4 except one participant who had an assistive life-space level of 1. All participants had a maximal life-space level greater than 3.

**Table 2.**
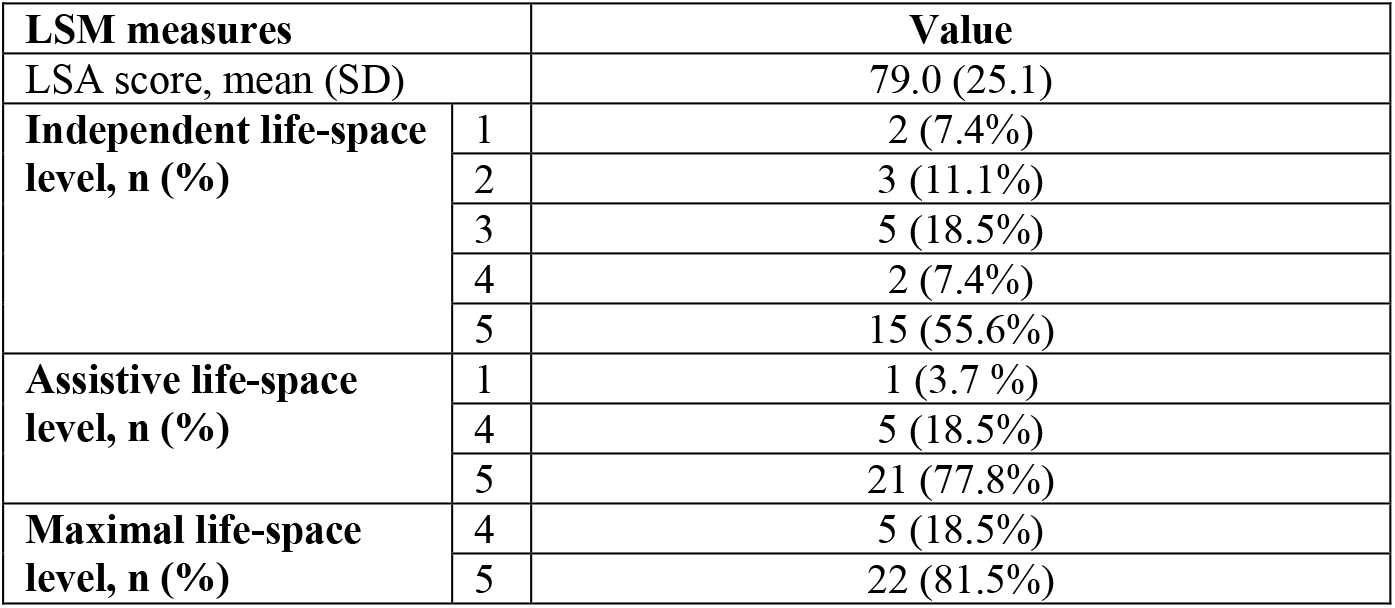
Summary of the LSM measures derived from the LSM questionnaire in our cohort.

| LSM measures |  | Value |
| --- | --- | --- |
| LSA score, mean (SD) |  | 79.0 (25.1) |
| Independent life-space level, n (%) | 1 | 2 (7.4%) |
|  | 2 | 3 (11.1%) |
|  | 3 | 5 (18.5%) |
|  | 4 | 2 (7.4%) |
|  | 5 | 15 (55.6%) |
| Assistive life-space level, n (%) | 1 | 1 (3.7 %) |
|  | 4 | 5 (18.5%) |
|  | 5 | 21 (77.8%) |
| Maximal life-space level, n (%) | 4 | 5 (18.5%) |
|  | 5 | 22 (81.5%) |

The descriptive characteristics of participants are shown in (Table 3). Mean age and mean SPPB score were 75 years and 9.4, respectively. Slightly more than half of the participants were male. The majority (92.6%) of the participants were white, and 85.2% of the participants had at least a college degree.

**Table 3.** Descriptive characteristics of participants.

| Characteristics | Value (n=27) |
| --- | --- |
| Age (years), mean (SD) | 75.0 (6.4) |
| **Gender, n (%)** |  |
| Male | 15 (55.6%) |
| Female | 12 (44.4%) |
| **Race, n (%)** |  |
| White | 25 (92.6%) |
| Black | 1 (3.7%) |
| Unknown | 1 (3.7%) |
| **Education, n (%)** |  |
| High School/Equivalent | 1 (3.7%) |
| College (13-16 years) | 13 (48.2%) |
| Post Graduate | 10 (37.0%) |
| Other | 3 (11.1%) |
| SPPB score, mean (SD) | 9.4 (2.0) |

The descriptive characteristics of the daily life-space mobility measures extracted from the GPS coordinates are shown in (Table 4). The medians of features measuring the distance and ellipse area deviated largely from the means. The mean excursion size for our participants was 83.1 km (one participant traveled out of state during follow up), while the median excursion size was 5.8 km. The average total distance travelled daily was 46.3 km and the median total distance travelled daily was 19.2 km. On average, our participants travelled outside home 2.3 times, had 2.3 clusters, and had a homestay rate of 0.5 every day.

**Table 4.** Descriptive characteristics of the extracted GPS features.

| Features | Min | Mean | SD | Median | Max |
| --- | --- | --- | --- | --- | --- |
| Excursion size (km) | 0.0 | 83.1 | 307.6 | 5.8 | 1645.1 |
| Excursion span (km) | 0.0 | 22.8 | 126.2 | 4.2 | 1718.7 |
| Total Distance (km) | 0.2 | 46.3 | 139.1 | 19.2 | 1836.8 |
| Ellipse Major Axis (km) | 0.0 | 29.1 | 168.6 | 6.0 | 2307.1 |
| Ellipse Minor Axis (km) | 0.0 | 4.4 | 8.3 | 1.5 | 44.7 |
| Ellipse Area (km<sup>2</sup>) | 0.0 | 657.8 | 5908.8 | 7.5 | 81032.3 |
| Number of Clusters | 1.0 | 2.3 | 1.3 | 2.0 | 7.0 |
| Entropy | 0.0 | 0.4 | 0.4 | 0.2 | 1.7 |
| Frequency of trips | 0.0 | 2.3 | 3.1 | 1.0 | 22.0 |
| Homestay | 0.0 | 0.5 | 0.4 | 0.7 | 1.0 |

(Table 5) provides the Spearman correlation coefficients along with the corresponding p-values between each GPS feature and each measure of LSM derived from the UAB-LSA questionnaire. All the GPS features were positively correlated with the composite LSA score except entropy, yet none were statistically significant. The excursion size was positively correlated with the maximum life-space level at a 0.05 significance level. The homestay rate was positively correlated with independent life-space level at a 0.05 significance level.

**Table 5.** Correlations between the GPS features and LSA measures.

| GPS Feature | Composite LSA score | Independent life-space level | Assistive life-space level | Maximum life-space level |
| --- | --- | --- | --- | --- |
| Excursion size (log) | 0.31 (0.110) | 0.16 (0.429) | 0.33 (0.091) | 0.40 (0.037) |
| Excursion span (log) | 0.22 (0.280) | 0.09 (0.662) | 0.27 (0.166) | 0.32 (0.106) |
| Total Distance (log) | 0.27 (0.181) | -0.05 (0.822) | 0.13 (0.524) | 0.22 (0.269) |
| Ellipse Major Axis (log) | 0.21 (0.283) | 0.12 (0.565) | 0.31 (0.119) | 0.35 (0.069) |
| Ellipse Minor Axis (log) | 0.18 (0.373) | -0.02 (0.911) | 0.20 (0.324) | 0.27 (0.174) |
| Ellipse Area (log) | 0.16 (0.419) | 0.02 (0.932) | 0.31 (0.119) | 0.37 (0.060) |
| Number of Clusters | 0.09 (0.653) | -0.00 (0.993) | -0.12 (0.542) | -0.01 (0.975) |
| Entropy | -0.04 (0.838) | -0.16 (0.414) | -0.25 (0.204) | -0.13 (0.503) |
| Frequency of trips | 0.23 (0.239) | -0.05 (0.806) | -0.19 (0.349) | -0.17 (0.390) |
| Homestay | 0.14 (0.476) | 0.38 (0.049) | 0.28 (0.154) | 0.18 (0.359) |

The p-value corresponding to each correlation coefficient is provided within parentheses.

(Figure 3) shows the relationship between each GPS feature and the composite LSA score separately for participants from UF and Yale. All the GPS features had a positive linear relationship with the composite LSA score except the homestay rate for participants from UF and Yale and entropy for participants from Yale. The magnitude of the linear relationship between GPS features and the composite LSA score were weakened for UF after combining data from two sites. Notably, the directionality of the correlations was flipped for entropy and homestay rate after combining the data.

**Figure 3.**
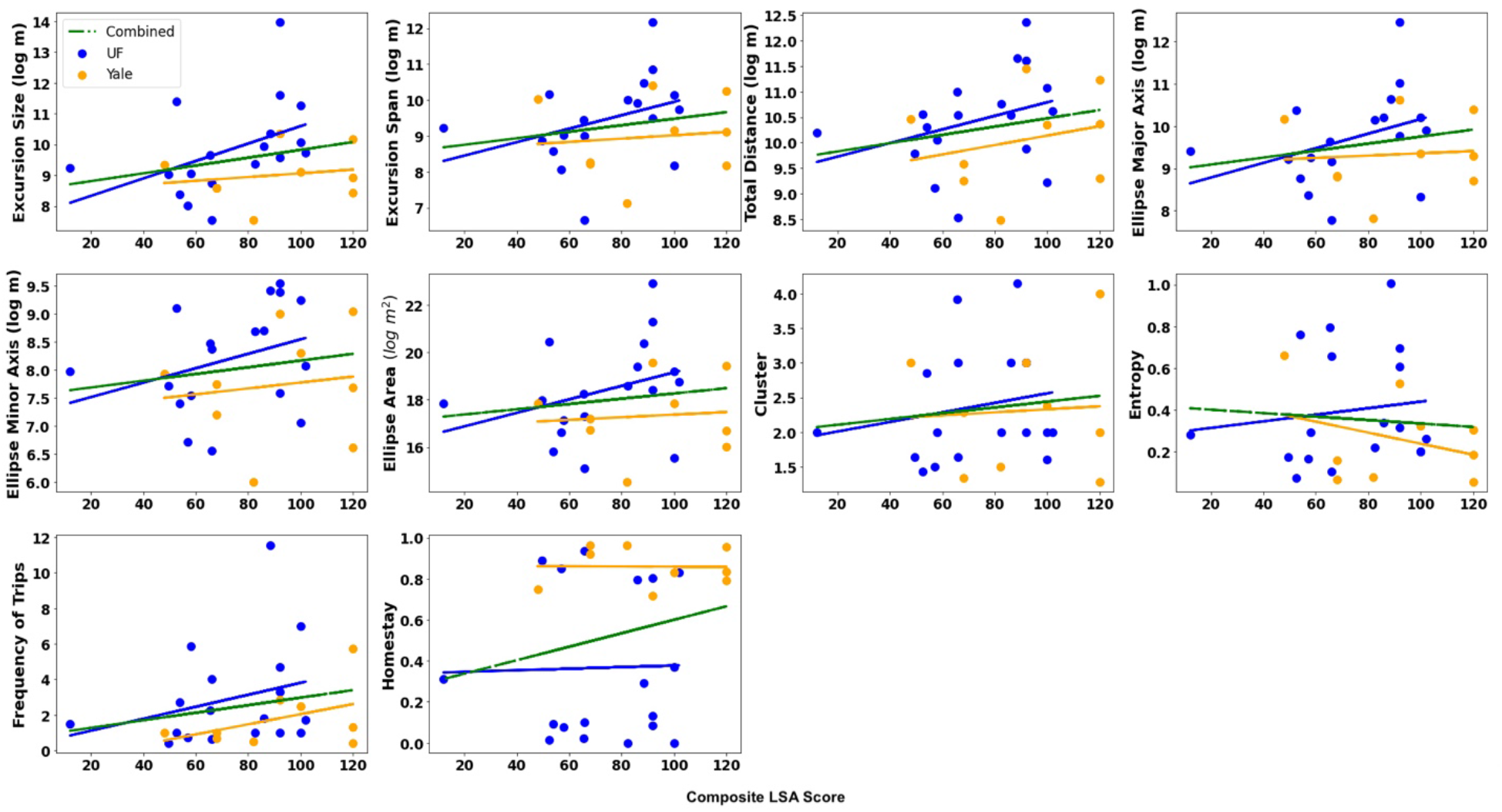
The relationship between each GPS feature and the LSA composite score. Each blue dot represents a participant recruited at the University of Florida site, and the blue line is the fitted regression line for the GPS feature and the LSA composite score. Each orange dot represents a participant recruited at the Yale University site, and the orange line is the fitted regression line for the GPS feature and the LSA composite score. The green line is the fitted regression line for the GPS feature combining participants recruited at the University of Florida and Yale University.

## Discussion

The goal of this study was to compare the LSM measures captured by GPS and LSM obtained from a self-reported questionnaire (UAB-LSA). We extracted ten GPS features collected using a smartwatch application to measure individual’s life-space mobility. Then, the correlation between four LSM measures extracted from UAB-LSA questionnaire and GPS features was examined. All the GPS features were positively correlated with the composite LSA score, except the entropy feature, yet none were significant.

Despite the ubiquity of UAB-LSA questionnaire in measuring LSM, it has limitations. The response of the questionnaire is restricted to the subjective interpretation of boundaries of life-space and depends heavily on the recall of the participants, which is especially problematic for older adults who may have trouble recalling events during the past month^9^. Zhu et al.^36^ also found UAB-LSA is susceptible to ceiling effect, which may be due to its poor discrimination of large life spaces. On the contrary, smartwatches equipped with a GPS receiver can continuously measure the LSM objectively and relieve participants of the burden of reporting their data. Additionally, the combination of the GPS features can provide a holistic view of individuals’ life-space pattern^27^, which is important in examining their LSM.

The relatively low correlation between GPS features and the composite LSA score may be caused by several reasons. First, unlike the self-reported questionnaires (e.g., UAB-LSA), GPS features measure more detailed LSM of an individual from day to day. For example, it enables measurement of the maximal distance the individual traveled (excursion size), the number of trips outside of home (frequency of trips), and variability of the locations they traveled (number of clusters) within a certain time frame. In contrast, the UAB-LSA instrument only measures the life-space on a scale of 1-5. Second, LSM questionnaires were filled out based on individuals’ recall of the previous four weeks, which may not reflect individuals’ actual LSM. This could result in discordance between individuals’ actual behavior and self-reported measures, as have been reported by a study examining the hypothetical functional capability and actual functional performance in older adults^37^. Relatedly, the results may present a true difference in LSM measured by a questionnaire versus LSM measured using GPS features. It is possible that the GPS measurements capture unique elements of day-to-day movement that cannot be reproduced using self-reported instruments. Our results are in-line with previous studies^26,36^ that have also shown limited convergent validity between GPS features and LSA scores.

The correlations between GPS features and maximal life-space levels were stronger than those for the other LSA measures. The main difference between these levels is the type of assistance reported. Although smartwatches have the capabilities of capturing high resolution GPS data that can better describe LSM, they fail to provide any information about the utilization of assistance, either by equipment or person. Apparently, the GPS features show stronger correlation when the individual’s needs for assistance are not accounted.

The results differed between participants from UF and Yale. Participants from Yale have higher composite LSA scores than participants from UF, as shown in (Figure 3), and all the participants from Yale obtained the highest independent life-space level. However, they showed relatively shorter distance in traveling, fewer trips outside home, and higher homestay rate. The reason for these differences is not entirely clear. Differences in regional restrictions due to the COVID-19 pandemic is a possible explanation, as participants from Yale were mostly followed between May and July in 2021. This data discrepancy also explains the positive correlations between the homestay rate and the LSM measures extracted from UAB-LSA questionnaire shown in this study.

Our study had limitations. First, the home addresses of participants in Yale were estimated. Nevertheless, accurate estimation of home addresses from GPS data has a twofold benefit: it protects individuals’ privacy and it is more flexible and accurate as some individuals may live somewhere different than their reported home, which could cause overestimation of LSM. Secondly, the participants in our study have a limited span of assistive and maximal life-space levels and distribution of their living space (in rural or urban areas) which may not generalize well to other populations. Finally, power to detect statistically significant correlations may have been limited given the relatively modest sample size.

## Conclusion

In summary, GPS features and self-reported questionnaires capture different dimensions of life-space mobility. Smartwatches equipped with a GPS receiver offer a unique opportunity to quantify the life-space mobility of older adults momentarily and provide high-resolution data than traditional life-space assessment instruments.

## Data Availability

All data produced in the present study are available upon reasonable request to the authors.

## Acknowledgements

This study was funded by the National Institute on Aging Claude D. Pepper Older American Independence Center NIH/NIA U24AG059624 and the National Institute on Aging P30AG021342 (Gill, Hajduk). Dr. Smail was funded on the T32AG062728 Translational research on aging and mobility (TRAM) program.

